# Convalescent Plasma in treatment of COVID-19: A review of evidence for a living systematic benefit-risk assessment

**DOI:** 10.1101/2020.08.24.20180729

**Authors:** Miranda Davies, Samantha Lane, Alison Evans, Jacqueline Denyer, Sandeep Dhanda, Debabrata Roy, Vicki Osborne, Saad Shakir

## Abstract

**Objectives:** We aimed to review the evidence for a living systematic benefit risk assessment for convalescent plasma use amongst patients with severe COVID-19 disease, based on currently available data.

**Methods:** The assessment used the Benefit-Risk Action Team (BRAT) framework. Convalescent plasma treatment in severe COVID-19 was compared to standard of care, placebo or other treatments. A literature search was conducted to identify published papers from January 1^st^, 2019 until July 8^th^, 2020. A value tree was constructed which included ranked key benefits and risks.

**Results:** We screened 396 papers from PubMed and 127 papers from Embase. Four studies were eligible for inclusion as they contained comparative data. Results from a randomised controlled trial revealed a non-statistically significant shortening of time to clinical improvement of 2.15 days (95% CI, −5.28 to 0.99 days) in the intervention group compared with the control group, with a possible signal of increased efficacy amongst a small subset of patients with severe disease (but not life threatening disease), however this study may have been underpowered. Interpretation of findings amongst the three controlled non-randomised studies were limited by small patient numbers, lack of randomisation, and confounding by co-administration of other treatments. Limited data availability at the current time precluded construction of a data summary table and further quantitative analysis.

**Conclusions:** There was insufficient evidence from controlled studies to complete a data summary table for a systematic benefit-risk assessment of the use of CP for severe COVID-19 disease at the current time, and as such a benefit-risk conclusion could not be made. Whilst uncontrolled case series have suggested positive findings with CP, results from these studies are very difficult to interpret. We provide a framework which can be updated when further data that have an impact on the benefit-risk become available.

**Article Summary:** Strengths and limitations of this study

- provides a living systematic benefit risk assessment based on currently available data for the use of convalescent plasma in patients with severe COVID-19 disease
- establishes a framework inclusive of ranked key benefits and risks for convalescent plasma in severe COVID-19 disease, into which additional data can be added as this becomes available facilitating re-assessment of the benefit risk profile
- uses a transparent framework (BRAT framework) which can be applied to other potential treatment options in this disease context
- insufficient data available at the current time from comparative studies to form a benefit risk conclusion

## 1 Introduction

The novel coronavirus subsequently called Severe Acute Respiratory Syndrome coronavirus 2 (SARS-CoV-2) which causes coronavirus disease (COVID-19) led to the World Health Organisation declaring a pandemic in March 2020 [1]. Whilst the majority of cases of COVID-19 are mild, there is an increase in the infection mortality ratio with increasing age, with the highest mortality risk amongst people aged ≥80 years [2]. Since the start of the pandemic, intensive worldwide scientific research has been conducted in a bid to identify safe, effective treatment options as quickly as possible. So far, remdesivir is the first treatment for COVID-19 to be recommended for authorisation in the European Union [3]. The European Medicines Agency is currently reviewing results from the RECOVERY study arm that involved the use of dexamethasone in the treatment of patients with COVID-19 admitted to hospital [4].

Convalescent plasma (CP) treatment, in which plasma containing antibodies is collected from patients who have recovered from COVID-19, is another treatment option currently being investigated. Two systematic reviews and exploratory meta-analyses included results for patients with respiratory infections of viral aetiology such as SARS-CoV and influenza virus who received CP [5, 6] and one of these included four studies inclusive of patients with SARS-CoV-2 [6]. Results suggested that CP may reduce mortality and would appear to be well tolerated with larger treatment effects if commenced early after symptom onset [5, 6].

In the US, on March 24^th^, 2020, the Food and Drug Administration (FDA) announced an emergency Investigational New Drug (IND) process to allow individual physicians to treat patients with serious COVID-19 disease with CP. On April 7^th^, 2020, the FDA announced a National Expanded Access Protocol administered through the Mayo Clinic. The protocol enabled a wider range of adults to be treated with CP by including those at risk of severe disease as well as those already suffering from severe disease [7]. On April 8^th^ 2020, the European Commission recommended that transfusion of COVID-19 CP, as an immediately available experimental therapy with low risk, should be considered as an urgent priority and its outcome monitored [8]. In addition to access to this treatment through compassionate use programmes, there are currently multiple RCTs being conducted across the world; as of June 30^th^ 2020, there were 113 studies involving CP, including expanded access programmes, registered on the clinical trials.gov website [9]. In the UK, the National Health Service Blood and Transplant (NHSBT) service is supporting two major clinical trials, RECOVERY and REMAP-CAP [10]. In the REMAP-CAP trial, CP treatment is indicated for people who have been in intensive care for less than 48 hours and have tested positive for COVID-19, whilst in the RECOVERY trial the effectiveness of CP is examined for patients with COVID-19 who are in hospital, but not in intensive care [10].

To date two systematic reviews have been conducted to assess the evidence in support of the use of CP in COVID-19. Rajendran and colleagues evaluated the effectiveness of CP transfusion (CPT) therapy in COVID-19 patients and concluded that in addition to antiviral/antimicrobial drugs, CPT could be an effective therapeutic option [11]. This review included five studies inclusive of 27 patients. More recently a living Cochrane systematic review by Valk and colleagues (published May 14^th^ 2020) aimed to assess effectiveness and safety of CP or hyperimmune immunoglobulin transfusion in COVID-19 [12]. Valk et al concluded that there was very low-certainty evidence on the effectiveness and safety of CP therapy for people with COVID-19 based on the data available at that time. However, a systematic benefit-risk assessment on the use of CP for COVID-19 treatment, based on currently available evidence, has not yet been conducted.

The Benefit-Risk Action Team (BRAT) uses a structured, descriptive framework to outline the key benefits and risks of a medication within a defined disease context. If sufficient relevant data are available, additional quantitative assessment can be used to further examine the benefit-risk profile [13]. Importantly, the framework improves transparency and consistency in the decision-making process and allows the robustness of any assumptions used in the assessment to be further explored using sensitivity analyses [14]. The method is dynamic, meaning that further data can be easily incorporated into the platform as this becomes available, and has been designed to assist communication with regulatory authorities [15], in addition to clinicians and public health bodies in the decision-making process on treatments for COVID-19. Benefit-risk assessments using this methodology in the context of severe COVID-19 disease have previously been applied to the anti-viral drug remdesivir [16] and the protease inhibitors lopinavir/ritonavir [17].

## 2 Objectives

The main objective of this study was to examine the benefit-risk profile of CP in COVID-19 patients compared to standard of care, placebo or other treatments. A key objective of this study was to provide a platform for a living systematic benefit-risk evaluation. Although initially this evaluation is likely to contain limited information, it is required due to the urgent unmet public need. Importantly it allows additional data to be incorporated as it becomes available, and re-evaluation of the benefit-risk profile.

## 3 Methods

### 3.1 Benefit-Risk framework

The BRAT framework was used to assess the overall benefit-risk of using CP as a treatment for COVID-19 compared to standard of care, placebo or other treatments. BRAT uses a six-step iterative process to support the decision and communication of a benefit-risk assessment: define decision context, identify outcomes, identify data sources, customise framework, assess outcome importance, and display and interpret key Benefit-Risk metrics [15]. Multiple settings for the use of CP are being investigated including high-risk individuals for prophylaxis, patients with mild disease, those with serious but not critical disease, intensive care unit (ICU) patients, or children. This assessment focuses on CP for the treatment of severe COVID-19 disease, which we defined as hospitalisation as a result of COVID-19 infection.

#### 3.1.1 Population of interest

Patients with severe COVID-19 who were treated with CP were the population of interest, while patients receiving standard of care, placebo or other treatments were the comparator of interest. Where standard of care was reported as the comparator group, this refers to no use of a specific pharmaceutical treatment for COVID-19 but provision of standard supportive therapy within the hospital setting.

#### 3.1.2 Identification of outcomes of interest – the value tree

Key benefits and risks were selected based on clinician judgement, i.e. those considered to be of clinical importance or potentially serious, and a visual representation of those key outcomes have been provided in a value tree. Key benefits and risks associated with the use of CP were identified for inclusion in the value tree from all available data sources, including the product information, regulatory assessment reports, and published literature. Key benefits (clinical endpoints) were derived from both published literature and in the case of ongoing studies, available clinical trial protocols. These benefits and risks, as presented in the value tree, were derived from both qualitative and quantitative data, and ranked according to perceived importance (benefits) and potential seriousness (risks). Two clinicians performed this ranking after discussion of the clinical significance of each benefit and risk.

#### 3.1.3 Data sources and customisation of the framework

A literature search was performed in PubMed and Embase using the following search strategy:

((plasma OR “blood plasma” OR “convalescent plasma”) AND (covid* OR SARS-CoV-2 OR (n-CoV OR nCoV) OR coronavirus)). Manuscripts were reviewed for eligibility and a clinical reviewer made the final decision on inclusion. Results were restricted to English language only (abstracts in English language were acceptable where sufficient data provided) and peer-reviewed publications since January 1^st^, 2019 to July 8^th^, 2020.

### 3.2 Outcome assessment

Where sufficient numerical data is available, a summary benefit-risk table allows visualisation of the magnitude of each benefit and risk. Risk differences and corresponding 95% confidence intervals (CI) are calculated for each outcome where both numerator (number of events) and denominator (number of patients at risk) are available for both the treatment group (CP) and comparator group. Since there was insufficient evidence for use of CP in COVID-19, quantitative outcome assessment was not undertaken.

#### Patient and Public Involvement (PPI) statement

There were no funds or time allocated for PPI so we were unable to involve patients.

## 4.0 Results

From literature searching we identified 396 papers from PubMed and 127 papers from Embase for CP. All papers were reviewed for information on benefits and risks relating to CP, including both qualitative and quantitative data. After initial review and removal of duplicates, 86 papers were identified with information on safety of CP and 19 papers with efficacy information for CP. These papers were reviewed further to determine whether they contained comparative data; only four papers compared CP to standard of care, placebo or other treatments.

Figure 1 displays the value tree of the key benefits and risks related to CP treatment in COVID-19. It is acknowledged that certain outcomes, such as mortality risk, could be considered as either a benefit (reduction in mortality risk) or a risk (increase in mortality risk) of CP treatment. As mortality risk is an important clinical endpoint in many COVID-19 trials, for the purposes of this assessment mortality risk has been considered under potential benefits. Risks have been classified as known risks and theoretical risks. The evidence for theoretical risks has arisen from knowledge of adverse effects observed with other viral diseases, including influenza, respiratory syncytial virus and SARS-CoV.

**Fig 1.**
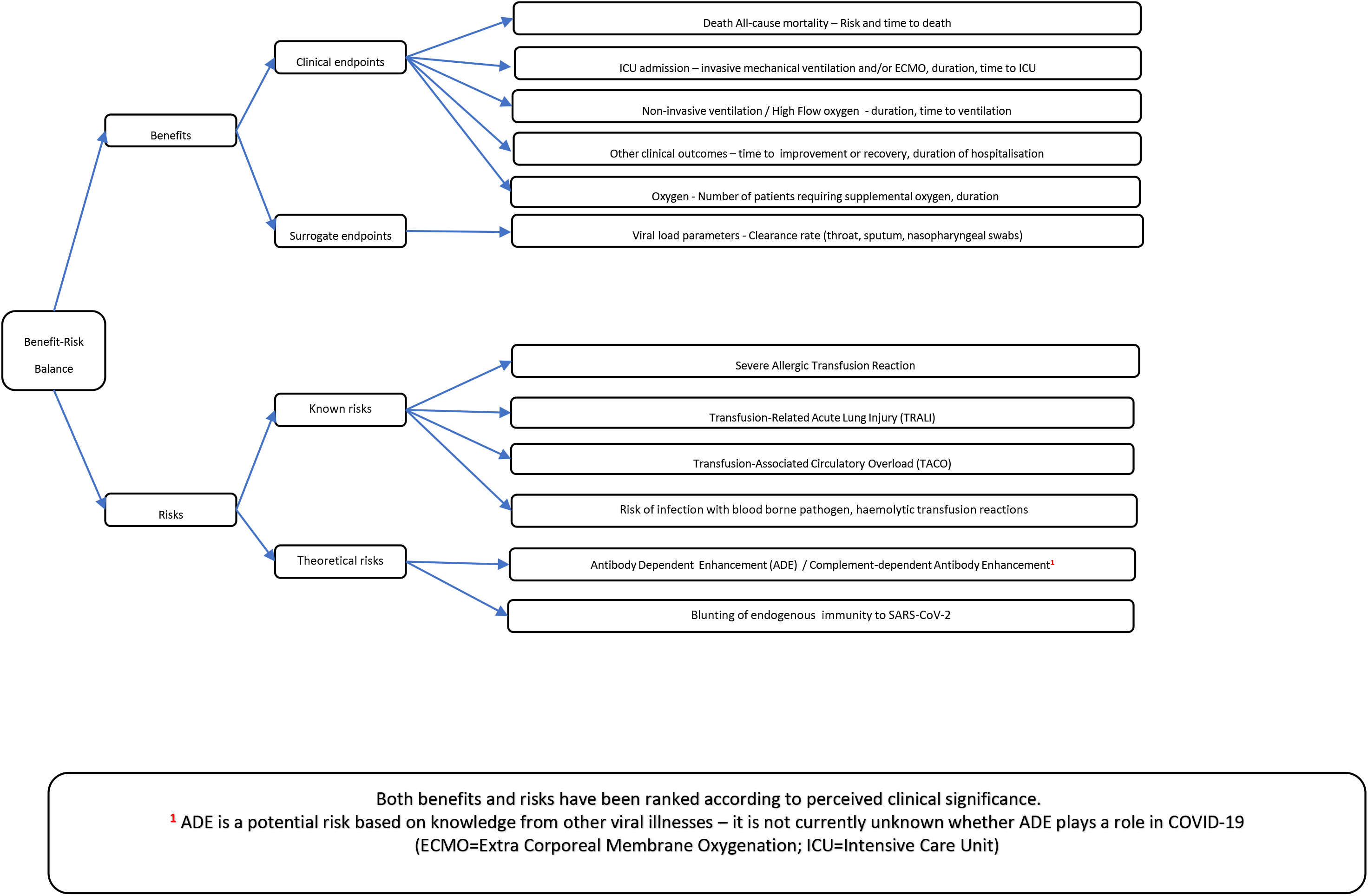
Value tree of key benefits and risks identified for convalescent plasma, ranked by order of clinical significance.

### 4.1 Benefits

Key benefits have been listed in the value tree in descending order of perceived clinical importance. Multiple case series have published findings following the use of CP amongst patients with COVID-19 [18-23]. Findings from these would appear to be encouraging, however interpretation is limited by the lack of a comparator group, the small number of patients, and confounding by co-administration of other COVID-19 treatments. As per the methodology for this benefit-risk assessment, only studies which included a control group were evaluated further. At the current time, we identified four comparative studies.

The first was an open label RCT conducted by Li et al *(published June 3^rd^ 2020)* which included 103 participants with laboratory confirmed COVID-19 that was severe (respiratory distress and /or hypoxemia) or life-threatening (shock, organ failure, or requiring mechanical ventilation) [24]. Two treatment arms consisted of patients treated with CP plus standard treatment (n=52) vs. standard treatment alone (control) (n=51). The primary outcome was time to clinical improvement within 28 days, defined as patient discharged alive or reduction of two points on a 6-point disease severity scale. Secondary outcomes included 28-day mortality, time to discharge, and the rate at which viral polymerase chain reaction (PCR) results turned from positive at baseline to negative at up to 72 hours. The target sample size was 200 patients; however the trial was terminated early due to a lack of patients and may have been underpowered. Study findings overall showed a non-statistically significant shortening of time to clinical improvement of 2.15 days (95% CI, −5.28 to 0.99 days) in the intervention group compared with the control group. Overall, clinical improvement at 28 days occurred in 27 patients (51.9%) in the intervention group vs. 22 patients (43.1%) in the control group (difference, 8.8%; 95% CI, −10.4% to 28%).

Amongst patients with severe disease, time to clinical improvement within 28 days was 4.94 days shorter (95% CI, −9.33 to −0.54 days) in the intervention group compared with the control group, and clinical improvement at 28 days occurred in 21 patients (91.3%) in the intervention group vs. 15 patients (68.2%) in the control group (difference, 23.1% (−3.9% to 50.2%). This is in contrast to patients with life threatening disease, in which there were no significant differences in the primary outcome or rates of clinical improvement at 28 days. Overall, no statistically significant differences were observed for any of the secondary clinical endpoints, however statistically significant differences were seen for rate of viral PCR conversion at up to 72 hours [viral nucleic acid negative rate at 72h (intervention group, 41/47(87.2%) control group, 15/40 (37.5%), difference 49.7% (32.0% to 67.5%) Odds Ratio (OR), 11.39 (3.91-33.18) p<.001].

More recently *(published June 19^tth^ 2020)*, Hegerova and colleagues reported the findings from a controlled non-randomised study in which results for 20 hospitalized patients treated with CP were compared to 20 matched controls with severe or life-threatening COVID-19 infection [25]. Patients’ clinical status was assessed using the WHO ordinal scale, and results suggested that clinical status at 14 days follow up was similar in both groups. A similar number of patients in each group were discharged home at 14 days (45%), whilst results showed a lower mortality risk at 14 days amongst the CP patients (2/20, 10%) compared to controls (6/20, 30%). No adverse events were reported in the CP group, although the risk in both groups of venous thrombosis at 14 days follow up was 20% [25]. Additional therapies for COVID-19 were also received by patients in both groups.

A further observational study conducted by Zeng et al *(published April 28^th^ 2020)*, retrospectively analysed the clinical outcomes of 21 patients who required ICU admission, of whom six patients received CP treatment, and compared clinical outcomes to the 15 patients who did not receive CP. The findings from this study suggested that all six patients who received CP achieved viral clearance compared to a quarter of patients who did not receive CP [4/15, 26.7%], however fatality rates were similar in both groups of patients [83.3%, 93.3%, p=0.5]. The authors state that failure of CP to reduce the mortality may be attributed to the late transfusion of CP (median duration of viral shedding before treatment was 21.5 days) [26].

The fourth comparative study by Duan et al *(published April 6^th^ 2020)* was a prospective non-randomised study in which outcomes were compared between ten patients with critical COVID-19 who received convalescent plasma (plus standard therapy) and ten historic control patients [27]. The historic controls were matched for age, gender and severity of disease, and received standard therapy. The primary endpoint was the safety of CP transfusion, whilst secondary endpoints included the improvement of clinical symptoms and laboratory and radiological parameters within three days after CP transfusion. Outcome data presented in a supplementary appendix revealed no reported deaths amongst patients who received CP compared to three deaths reported amongst control patients (30%) [28]. Favourable results were observed with respect to the proportion of patients with improvement in clinical status and the proportion discharged following CP compared to controls. However, limitations to interpretation of results included the small sample size and lack of adjustment for additional confounding factors. Furthermore, the period of follow up during which outcomes, including mortality, were assessed was limited to three days post transfusion whilst the follow up period was not clear amongst control patients. Therefore follow up period was short, and it is not known whether follow up periods for both groups were comparable. Whilst no serious adverse reactions or safety events were recorded after CP transfusion, AEs were not reported for the control group.

### 4.2 Risks

The key risks identified in the value tree were identified from a range of data sources and ranked according to perceived seriousness. The incidence of the transfusion reactions included in the value tree have been described in an unpublished report by the US FDA Expanded Access Program for COVID-19, based on the first 20,000 hospitalized patients transfused with CP [29]. Treating physicians evaluated relatedness, and transfusion-related serious adverse events (SAEs) were independently adjudicated. One hundred and forty-six SAEs classified as transfusion reactions were reported (<1% of all transfusions). Transfusion related reactions included transfusion associated circulatory overload (TACO), transfusion-related acute lung injury (TRALI), and severe allergic transfusion reactions. The report also classified mortality that occurred within four hours of transfusion as a transfusion associated reaction. Amongst transfusion related reactions, TACO was reported at an incidence of 0.18% (95% CI 0.13%, 0.25%) followed by severe allergic transfusion reaction (0.13% (95% CI 0.09%, 0.19%)), and TRALI (0.10% (95% CI 0.06%, 0.15%)). Mortality within four hours of transfusion that was assessed as related was reported in 13 patients (0.06% (95% CI 0.04%, 0.11%)) [29].

This study also described the incidence of thrombotic and cardiac events. Within seven days of completion of the COVID-19 CP transfusion, 1,136 other SAEs were reported. Of these SAEs, 87 thromboembolic or thrombotic events were reported, 406 sustained hypotensive events requiring intravenous pressor support were reported, and 643 patients suffered a cardiac event. Notably, the vast majority of the thromboembolic or thrombotic complications (n=55) and cardiac events (n=569) were judged to be unrelated to the plasma transfusion. The authors state that in aggregate, adverse cardiac events occurred in ~3% of patients transfused with COVID-19 CP. The vast majority of adverse cardiac events of interest (88%) were deemed unrelated to the plasma transfusion by the treating physicians. Overall, there was insufficient evidence to suggest that thrombotic and cardiac events were potential risks associated with CP and hence neither of these events have been included in the value tree. However, it is acknowledged that plasma naturally contains procoagulants [30] and the effects of these in the context of COVID-19 disease with respect to risk of thrombotic events is currently not known.

There are additional risks associated with transfusions including infectious disease transmission, however this has been much reduced in recent decades due to donor screening and testing [31], hence this risk is considered to be low, but may vary between countries. The risk of haemolytic transfusion reactions is similarly considered to be low, as transfusion services provide blood group ABO compatible fresh frozen plasma to patients [31].

An additional potential risk of CP transfusion includes the development of antibody-dependent enhancement (ADE), which has been previously recognised in other viral illnesses [32, 33]. It is characterized as either the facilitation of viral entry into cells by antibody or the enhancement by antibody of viral toxicity [34]. A further related risk is complement-dependent antibody enhancement, which has also been seen in other viral illnesses such as Ebola, and it is not yet known whether this plays a role in COVID-19 [34]. Theoretically the administration of antibodies to those exposed to SARS-CoV-2 may prevent COVID-19 disease in a manner that reduces the immune response, possibly leaving such individuals vulnerable to subsequent reinfection [35].

In the RCT by Li et al, two participants reported transfusion-related adverse events following CP transfusion. One patient in the severe COVID-19 group developed a non-severe allergic transfusion reaction and also a probable non-severe febrile non-haemolytic transfusion reaction. The other patient, who was in the life-threatening COVID-19 group, developed possible severe transfusion-associated dyspnoea. No SAEs were reported in the three controlled non-randomised studies [25-27].

### 4.3 Quantitative data

Due to the lack of available efficacy and safety data available from comparative studies it was not possible to perform further quantitative analysis.

## 5 Discussion

The primary objective of this study was to perform a living systematic benefit-risk assessment of the use of CP for the treatment of severe COVID-19 disease, using methodology previously published [13, 16, 17]. An integral part of this process was the identification of key benefits and risks from all available data sources potentially associated with CP, within this disease context. At the current time, the availability of data from comparative studies pertaining to these key outcomes (as per the value tree) was limited, which precluded further quantitative analysis. However, the identification of key benefits and risks within this framework allows the incorporation of further data relating to these outcomes to be incorporated as and when this becomes available.

Whilst a systematic benefit-risk assessment could not be undertaken at the current time due to the lack of data, we discuss the benefit and risk data available to date. Whilst uncontrolled case series have suggested positive findings with CP, results from these studies are very difficult to interpret, given the limitations previously discussed. We identified only four controlled studies, inclusive of one RCT and three controlled non-randomised studies.

In the RCT, findings for the primary outcome of time to clinical improvement amongst the 103 patients included were not statistically significant, however it is acknowledged that the study may have been underpowered due to its early termination. It is of note that a statistically significant result for this outcome was obtained when the intervention group was restricted to patients with severe disease only (but not life-threatening disease) compared to the control group. This may reflect the administration of this therapy earlier in the disease process, when it is considered that antibody-based therapies work best [36, 37].

Nevertheless, the overall median time between onset of symptoms and randomisation was 30 days, and the authors state that it is unclear whether earlier treatment would have resulted in greater benefit [24]. The question of whether earlier treatment may have resulted in greater benefit also applies to study findings by Zeng et al, in which the median days to treatment was 21.5 days [26]. In the study by Hegerova et al, the median time from hospitalization to CP was earlier at 2 days (IQR 1-4.3), however the study does not report on the median time from onset of symptoms [25]. The mean time from onset of illness to CP transfusion amongst the ten patients who received CP in the study by Duan et al was 16.5 days [27].

Based on these studies, evidence of benefit at the current time is limited. The results from the study by Li et al may suggest some shortening of the time to clinical improvement amongst patients with severe disease, but this possible signal is based on a small subset of patients (n=23) and would require further investigation in adequately powered randomised controlled studies. However statistically significantly higher viral clearance rates seen at 24, 48 and 72 hours were observed amongst patients who received CP, which would suggest that viral clearance may be achieved before clinical improvement is observed [24]. Interpretation of study findings from the three controlled non-randomised studies is limited by the lack of randomization, small patient numbers involved, and confounding by additional risk factors including co-administration of other COVID-19 treatments.

The value tree identified potential risks associated with CP. TACO is considered to be one of the most common serious adverse effects of transfusion [34] and it is possible that elderly patients with acute lung injury may be at increased susceptibility [34]. Overall, limited safety data was available from the four controlled studies identified. Two patients experienced transfusion-related adverse events in the study by Li et al (3.8%) and both patients recovered following supportive treatment [24]. This figure would appear to be consistent with reported incidence rates of TACO and TRALI previously published, in which active surveillance estimates of TRALI were ~0.1% of transfused patients but up to 5% to 8% in groups of intensive care unit populations [38]. The authors of the study by Hegerova et al stated there did not appear to be an increased risk of venous thromboembolism in CP patients, although the incidence was high in both groups despite heparin prophylaxis (20% at 14 days post follow up) [25].

An additional potential risk of CP transfusion is the development of ADE which has been previously recognised in other viral illnesses [32, 33]. A further related risk is complement-dependent antibody enhancement, which has also been seen in other viral illnesses such as Ebola, and it is not yet known whether these play a role in COVID-19 [34].

A limitation of this study was that there was insufficient data to produce a data summary table, or to perform any quantitative analysis, therefore we were unable to complete all stages included in the BRAT methodology. The only RCT included in this assessment was probably underpowered to detect the primary outcome of time to improvement, and overall showed no statistically significant improvement amongst patients who received CP. There was also very little risk data amongst controlled study populations, and it acknowledged that the role of ADE in COVID-19 has yet to be investigated. However, we have presented a value tree inclusive of key benefits and risks which can be used to guide further assessments, as additional data becomes available. As per the eligibility criteria for BRAT methodology, only peer reviewed published papers have been included in this assessment, however it is acknowledged that further studies containing comparative data are currently available as pre-prints and therefore have not yet been peer reviewed. One of these studies is an RCT in which CP was compared to standard of care therapy amongst patients hospitalized for COVID-19 in the Netherlands [39]. The primary endpoint was day-60 mortality and key secondary endpoints included hospital stay and WHO 8-point disease severity scale improvement on day 15. This study was terminated prematurely after 86 patients had been enrolled (planned sample size was 426 patients). Patients were noted to have had COVID-19 related symptoms for a median of 10 days (IQR 6 – 15) and admitted for median of 2 days (IQR 1 – 3 days). Of the patients who had been enrolled, the majority for whom blood samples were available (n=66) already had high neutralizing antibody titres which were comparable to the 115 recovered donors screened for CP plasma donation. As a result of this, the decision was made to terminate the study, as it was considered very unlikely that the overall study population would benefit from CP therapy without a change in the patient recruitment strategy. No statistically significant differences in mortality (adjusted OR 0.95, 95% CI 0.20 – 4.67, p=0.95) or improvement in the day-15 disease severity (adjusted OR 1.30, 95% CI 0.52 – 3.32, p=0.58) was observed when the study was suspended.

A further non-randomised controlled study in the U.S by Liu et al compared outcomes of 39 hospitalized patients with severe to life-threatening COVID-19 who received CP transfusions with a cohort of retrospectively propensity score-matched controls [30]. Patients were evaluated for supplemental oxygen requirements and survival at days 1, 7, and 14 post-transfusion. The median duration of symptoms prior to initial presentation was 7 days (IQR 0-14) and the median time between admission and transfusion was 4 days (IQR 1-7) days. Matching of controls was performed on multiple factors including administration of hydroxychloroquine and azithromycin, intubation status and duration, length of hospital stay and oxygen requirement on the day of transfusion. Control patients were matched to plasma recipients by length of stay prior to transfusion. Improvement of clinical symptoms, as assessed by need for respiratory support at day 14, included 39 participants in the intervention group and 156 participants in the control group. Clinical condition had worsened in 18.0% of the plasma patients and 24.3% in the control patients (p=0.167). The adjusted odds ratio for worsening oxygenation on day 14 was 0.86 (95% CI: 0.75-0.98, p=0.028). On days 1 and 7, the plasma group also showed a reduction in the proportion of patients with worsened oxygenation status, but the group difference did not reach statistical significance. Overall, 12.8% of plasma recipients and 24.4% of the control patients died and 71.8% and 66.7% had been discharged, respectively. Overall improved survival was observed for the plasma group (log-rank test: p=0.039). In a Cox regression (time to event) analysis, CP transfusion was significantly associated with improved survival in non-intubated patients (adjusted hazard ratio (HR) 0.19, (95% CI: 0.05 −0.72),p=0.015), which was not observed in intubated patients (adjusted HR 1.24, 95% CI (0.33-4.67,p=0.752)). In this study, the authors found no evidence that the effect of CP depended on the duration of symptoms (p=0.19 for the plasma by duration interaction). No significant transfusion-related morbidity or mortality were observed in the CP recipient cohort. It is of note that results were adjusted for hydroxychloroquine and azithromycin, intubation status and duration, length of hospital stay, and oxygen requirement on the day of transfusion. However, results were not adjusted for age and gender, and the sample size was too small to allow for further sub-group analyses.

Complete data from these two studies will be included in subsequent CP benefit risk assessments, following their publication.

## Conclusion

There was insufficient evidence from controlled studies to complete a data summary table for a systematic benefit-risk assessment of the use of CP at the current time, and as such a benefit-risk conclusion could not be made. It is of note that whilst uncontrolled case series have suggested positive findings with CP, results from these studies are very difficult to interpret. We provide a framework in which anticipated key benefits and risks have been identified, which can be updated when further data that have an impact on the benefit-risk assessment become available.

## Data Availability

Data used in this analysis are available from the references supplied.

## Footnotes

### Contributorship statement

MD assisted with study design, literature searching, identified outcomes of interest, constructed the value tree and wrote the first draft of the manuscript. SD, DR, SL, JD and AE assisted with study design, literature searching and data extraction. VO and SAWS assisted with the concept, study design and manuscript revisions. All authors reviewed, contributed to revisions and approved the manuscript and accept full responsibility for its overall content.

### Data sharing statement

Data used in this analysis are available from the references supplied.

### Ethics approval

This study was conducted in accordance with international ethical guidelines. Ethics approval was not required for this study.

### Transparency statement

The lead author affirms that the manuscript is an honest, accurate, and transparent account of the study being reported; no important aspects of the study have been omitted. All authors reviewed, contributed to revisions and approved the manuscript and accept full responsibility for its overall content.

### Consent to participate

Not applicable

### Consent for publication

Not applicable

### Availability of data and material (data transparency)

Data used in this analysis are available from the references supplied.

### Code availability (software application or custom code)

Not applicable

### Patient consent

Not required

